# Machine learning methodology using a masked neural network for robust genetic risk score calculation from noisy and missing data

**DOI:** 10.64898/2026.05.18.25341725

**Authors:** Steven Squires, Michael N. Weedon, Richard A. Oram

## Abstract

**Purpose:** Genetic risk scores (GRSs) are summaries of genetic data that can improve prediction of disease risk and progression. GRSs are increasing available but rely on high quality input data to produce good output results; with noisy or missing inputs the GRS may be inaccurate. We aimed to develop a method to produce a robust estimate of the GRS when input data is missing, noisy or both.

**Approach:** We developed a neural network approach, named masked-MLP, for robust GRS calculation trained on a set of GRS scores calculated on clean data. The masked-MLP includes additional input data and has noise inserted during training, both which make the model more robust.

**Results:** A GRS for type 1 diabetes (T1D) calculated on input data with 10% of the data corrupted had a Spearman rank correlation to the clean GRS of 0.669 (0.665-0.674) while the equivalent for the masked-MLP was 0.951 (0.950-0.952). For the same data the area under the receiver operating characteristic curve for separation of T1D from population samples fell from 0.919 (0.904-0.932) to 0.808 (0.787-0.827) for the GRS while the masked-MLP fell to 0.910 (0.895-0.924).

**Conclusions:** The masked-MLP was more robust to noise when calculating a GRS than using standard approaches. Our approach has the potential to ensure both improved research and clinical outcomes due to more reliable GRS calculation.

## 1 Introduction

Genetic risk scores (GRSs), also known as polygenic risk scores, compress genetic information related to a trait or disease into a single number [1]. An effective GRS for disease provides discrimination power between cases and controls allowing prediction of onset and progression [2].

GRSs for a disease are typically generated by applying genome wide association studies (GWAS) to a dataset partitioned into those with and without the condition [3]. A GWAS assesses which single nucleotide polymorphisms (SNPs) are statistically significantly associated with the disease, along with the strength of association (the weights), and the risk varying allele. A standard GRS with *p* SNPS is calculated by multiplying the weight (*β*) of each statistically significant SNP by the number of effect alleles (*x*_*i*_); these are then summed to give the final score, 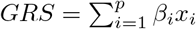.

There are also more complicated GRSs which, in addition to the standard linear part, also include non-linear combinations of SNPs. For example, pairwise interaction between SNPs can together alter the risk; these can become complicated especially if the combinations can be contradictory or exclusive [4]. Alternatively machine learning can be used to find more complex higher order functions between SNPs potentially improving discriminative power [5]. These more complicated, non-linear GRSs can produce improved predictive performance and are likely to become more common as datasets become larger, methods become more efficient and computer power increases.

Several software programs such as PLINK [6] can calculate a simple linear GRS if all the required SNPs are available and they are all accurately measured. More complicated non-linear GRSs can be more difficult to calculate even if all SNPs are available, requiring the use of more specialised programmes. However, if any SNPs are not available or if their quality is low then standard approaches will not produce the correct GRS. The purpose of this paper is to develop and demonstrate a methodology using machine learning that can produce an accurate GRS when SNPs are missing or inaccurately measured.

Genetic data, as relevant to GRS generation, can broadly be categorised into: small chips that generate a small number of SNPs, array data that produce hundreds of thousands of SNPs, and whole genome data which contain millions of SNPs. Whole genome sequencing is still expensive and does not currently scale. The other two methods often do not produce all the necessary SNPs to calculate a GRS.

In addition to missing SNPs the quality of the estimation of the values of those SNPs can be variable. If it known which SNPs are of poor quality they can be excluded, which produces the issue of missing data; alternatively if included they provide noisy input data to the GRS which will reduce the quality of the risk score.

If the aim is to investigate summary level data, such as for assessing the quality of a GRS on a particular population, the accuracy of individual samples may not be critical. Either a GRS can be calculated from noisy or missing data with an accepted level of error. Alternatively methods exist to generate GRSs from summary statistics [7] which can largely bypass issues of individual variation. However, neither of these methods can give a robust GRS on individual samples. In addition, if the noisy or missing inputs result in inaccurate scores then judgements about the GRSs on populations may be incorrect. For example, if the noise in the data results in poor discrimination between classes because of the quality of the SNP data, it might erroneously be concluded that the GRS is ineffective on that population.

Standard approaches to missing or poor quality data can be partially addressed by quality control (QC) and imputation. QC in this context is a set of procedures to remove SNPs that are of poor quality [8]. Imputation involves the use of panels of much larger numbers of SNPs from known populations which are compared to the available SNPs in the desired dataset and if possible missing data is inferred [9]. However, quality control tends to remove SNPs, resulting in more missing SNPs, and the assessment of which poorer quality SNPs to remove may not always be correct. Imputation is a statistical approach which replaces missing SNPs with those correlated with the original (a proxy) but the correlations are rarely perfect between the original and proxy SNP. The effect of QC and imputation is that SNPs can still be missing or noisy. Ideally there would be an approach that could take in the data and produce a robust GRS in the presence of noisy data.

Machine learning has had a large impact in multiple fields but its effect on GRSs has been more modest. For the problem of missing SNPs there may be some use of trained neural network models which can compensate for missing SNPs [10] although bespoke models need to be trained for each set of missing SNPs, and the ability to compensate largely depends on the correlations between SNPs in the GRS.

We propose, and demonstrate, a method of calculating a robust GRS using a machine learning approach. The inputs to our model include the original SNPs of the GRS along with additional SNPs correlated with those originals. During training we impose additional simulated corruption into the input data which forces the model to compensate for the extra noise. By doing so we produce a model which is robust both to noise (or corrupted SNPs) and to the replacement of the specific SNP with correlated alternatives. We primarily trained and tested the model using a non-linear GRS developed for type 1 diabetes (T1D) on a large dataset from UK Biobank (UKBB) [11]. We further tested the approach on two other T1D GRSs and a Celiac disease GRS to demonstrate its generalisability.

## 2 Methodology

### 2.1 Overall approach

A standard linear GRS is calculated by 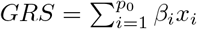 where *β*_*i*_ is the weight found from the GWAS, *p*_0_ is the number of SNPs in the GRS, and *x*_*i*_ is the allele count of 0, 1 or 2. More complicated GRSs might include non-linearities into the function, producing the GRS by some function *GRS* = *f* (**x**_0_), where 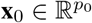 is the set of SNPs.

Our approach to develop a method for robust GRS calculation with noisy and missing input data utilises the original GRS calculation method to produce estimates on uncorrupted data. We then train a neural network model with those GRS estimates as labels for a regression task with the objective of finding *g*(**x**) ≈ *GRS*, where *g* is the function produced by the neural network and **x** is a set of SNPs, which can be different than the exact GRS SNPs.

We aim to accurately estimate the GRS even if there are missing or noisy SNPs. There are three key aspects of this method: the neural network architecture, additional input SNPs, and targeted corruption of the data during training.

### 2.2 Model and training

A schematic diagram for the model when it is running inference is shown in Figure 1. The input vector **x** ∈ ℝ^*p*^ (the SNPs) is fed into the trained model where the neural network encoder maps it to a lower dimensional latent space **z** ∈ ℝ^*r*^. The latent vector is then mapped to three outputs: the mask predictor 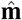, the estimate of the input vector 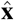 and the model’s estimate of the GRS score 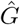. The mask prediction is the model’s estimate of which of the input data is corrupted (wrongly labelled) and the feature vector prediction is the model’s estimate of what a clean (not noisy) input vector would be.

**Figure 1:**
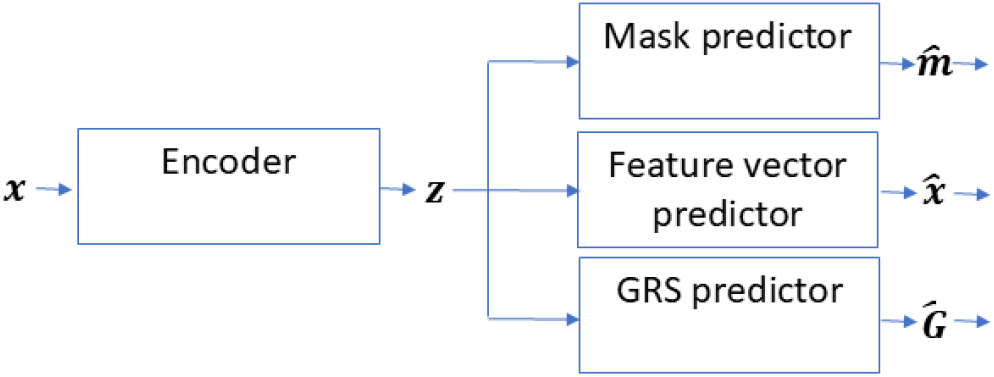
Schematic diagram of the model when performing inference. The model during inference consists of an encoder built as an MLP and linear mappings to the three outputs. The input data **x** consisting of *p* SNPs is fed into the model, the trained encoder produces a latent vector **z** which is then mapped to three outputs: the model estimate of the falsely labelled inputs 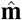, the model estimate of the true input vector 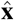 and the GRS estimate 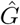.

The encoder is an multi-layer perceptron (MLP) with a *p*-dimensional input and two hidden layers with 50 and 30 neurons respectively and an output latent dimension, *r*, of 20 neurons. We use rectified linear units (ReLU) for the activation function of all the neurons. The mask, feature vector and GRS predictors are single layer mappings from the latent layer (with 20 neurons) to the required output. For the mask and feature vectors the output dimension is *p* while for the GRS prediction is 1. MLPs are universal function approximators [12] therefore should be able to find a function to map noisy inputs to the correct GRS output if it is possible.

The input to the model, **x** ∈ ℝ^*p*^, is an important part of our approach. The original GRS consists of *p*_0_ defined SNPs, but these can be unavailable or of poor quality. We utilise SNPs that are known to be correlated with the original SNP (called proxies) and include these in addition to the original. If the original is unavailable we replace it with the SNP with the highest correlation to the original that is available. Therefore the input **x** ∈ ℝ^*p*^ is the set of input SNPs which includes the original *p*_0_ required by the GRS with an additional 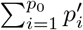 which is the sum of the proxy SNPs included with each original to give the total input as *p*. The choice of how many proxy SNPs to include for each original SNP is discussed in Section 2.3.

A schematic diagram of the model when it is being trained is shown in Figure 2; the differences from the model during inference (shown in Figure 1) is the additional of the mask and corruption generator which takes the input data **x** and artificially corrupts it before feeding it into the encoder. Once the model is fully trained the mask generator and corruption generator are removed and the model can be used for inference.

**Figure 2:**
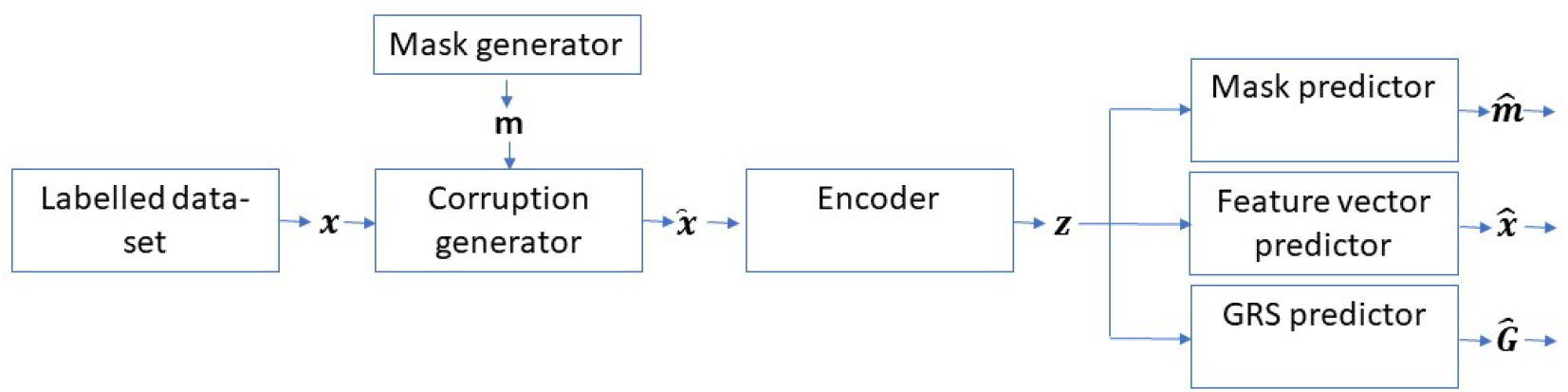
Schematic diagram of the model during training. The mask generator and corruption generator are added to the architecture during the training phase; together they corrupt the input vector **x** such that a noisy input, 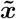 is fed to the encoder, forcing the model to learn a robust representation.

The purpose of the mask and corruption generators is to add targeted noise into the input data during training such that the model is forced to find a representation of the data in the latent vector **z** that is not overly dependent on individual SNPs and can still produce an accurate GRS prediction even with the corruption of those SNPs. The mask generator randomly assigns a mask to the input **x** with the probability of masking per-SNP being defined as a hyper-parameter (discussed in Section 2.3), those SNPs to be corrupted are marked with a 1 and uncorrupted with a 0. The corruption generator randomly changes the masked SNP to 0, 1, 2.

The labelled dataset 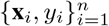 is made up of a set of *p*-dimensional vectors **x**_*i*_ each with an associated GRS label *y*_*i*_ for n-samples. During training, for one input sample, the mask generator produces a binary output that specifies whether a SNP is to be artificially corrupted, with 1 or 0 denoting SNPs to be corrupted or not, respectively. The mask, **m**_*i*_, is fed into the corruption generator along with the input data, **x**_*i*_. The corruption generator then alters those SNPs denoted by the mask to produce 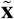, which is fed into the encoder which produces a latent vector **z**_*i*_. Three mappings are then produced which converts **z**_*i*_ into the predicted mask, 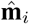, the predicted uncorrupted feature vector, 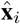, and the GRS prediction, 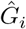.

A toy example is shown in Figure 3 for one input sample running through the model during training. The true input data is given by **x** with elements {2, 0, 1}, a mask is generated with instructions to corrupt the first two elements and a corrupted vector is generated. The input data is altered by the combination of mask and corrupted data giving the new corrupted input which is fed into the encoder which produces the **z** which is then mapped to the predicted mask, predicted feature vector and output GRS.

**Figure 3:**
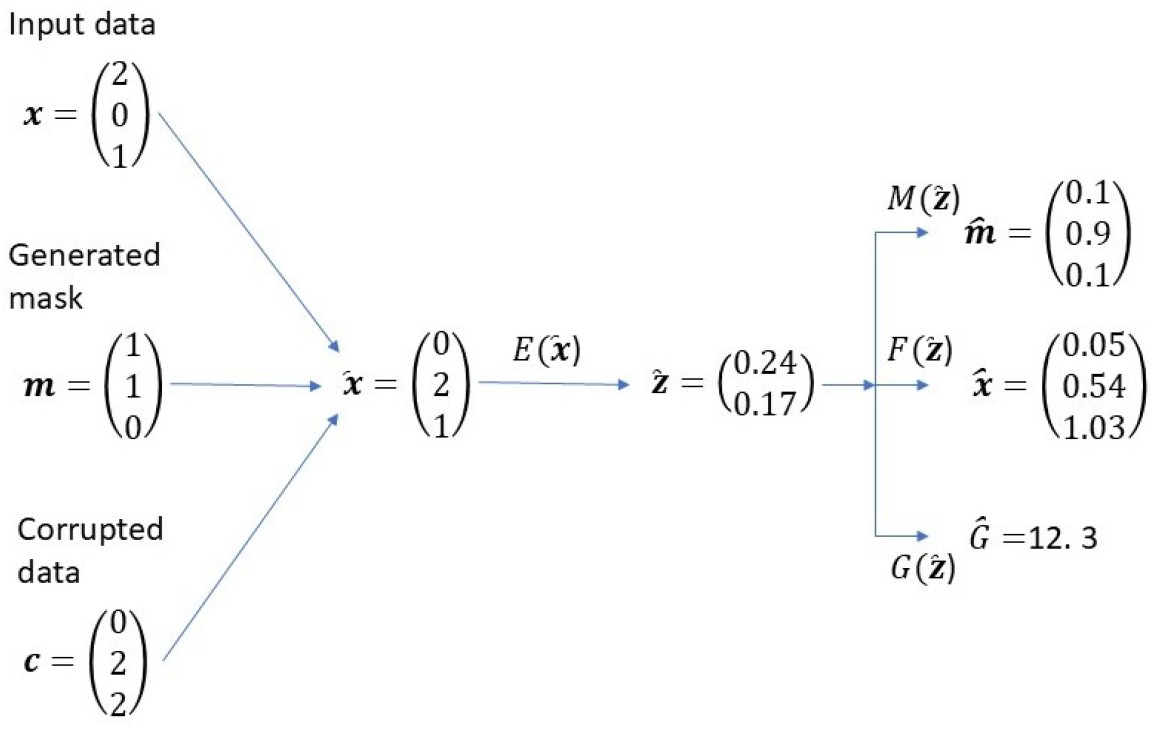
A toy example of one sample being run through the model during training. The input data is corrupted by the combination of the generated mask and corrupted data, which alters two of the input values to their corrupted versions. The encoder then maps that corrupted input to the latent vector followed by the three linear mappings to produce the three outputs.

For the full model the loss function (for one sample) is given by:

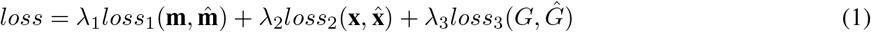

where *λ*_1/2/3_ are the weightings of the losses for the predicted outputs. *loss*_1_ is binary cross entropy, while *loss*_2_ and *loss*_3_ are both mean squared errors.

### 2.3 Model hyper-parameters and training details

The model was built using PyTorch [13] and trained end-to-end using the Adam optimiser [14]. We varied only the learning rate from the defaults, used a batchsize of 20, and saved the model parameters at any epoch if the validation error was lower than any previous epoch. We used *λ*_1/2/3_ = 1 (see Equation 1) but they could be adjusted to potentially improve performance.

An important hyperparameter is the number of additional correlated SNPs to include for each original SNP. While there are multiple potential approaches to take we pragmatically chose to select three additional SNPs for each original SNP; this is to demonstrate the effectiveness of the approach rather than an optimal method. We selected the additional SNPs by their level of correlation with the original without factoring in any additional information such as the SNP quality. There are alternatives, such as selecting the additional SNPs by assessing their quality in conjunction with the correlation between the SNP and the original.

Another hyperparameter is the masking probability. For our method we used a uniform probability, testing different levels during training. A masking probability of 0.2 during training means that every input SNP has a 20% change of being changed to one of 0, 1, 2, with equal probability of each. We trained multiple models with different masking probabilities ranging from 0 to 0.25 in increments of 0.05. For each masking probability we selected the final model based on the performance, as defined by the Spearman rank correlation coefficient to the labels, of the model on the validation set.

### 2.4 Testing model performance

To test the performance of our methodology we used the UK Biobank [11] dataset which is a prospective cohort dataset of approximately 500,000 people recruited from the United Kingdom to build a dataset to advance healthcare research. UK Biobank has ethical approval from the North West Multi-centre Research Ethics Committee and all participants gave their consent. Additional ethics approval is not required.

We considered two diseases: T1D with 387 cases and Celiac disease with 1,184 cases. The T1D cases were defined the same as previous work [4] as: clinical diagnosis under 21 years, on insulin within a year of diagnosis, at recruitment using insulin, not having type 2 diabetes and not on oral-antihyperglycemic agent. Celiac disease was defined by ICD10 reporting and self-reporting.

The UK Biobank genetic data we used consists of 487,409 samples imputed using Minimac [9] servers and a combination of the 1000G [15] and Haplotype Reference consortium panels [16]. For the T1D dataset we separated the 387 T1D cases into a test set. For the rest of the data we randomly partitioned it into 70% training (340,915), 15% validation (73,053) and 15% testing (73,054) sets. For the Celiac disease dataset, similarly we separated the 1,184 cases into a test set. The rest of the data was randomly partitioned into 70% training (340,357), 15% validation (72,933) and 15% testing set (72,935).

We considered three GRSs for T1D and one for Celiac disease. To demonstrate our method we focused on a T1D GRS consisting of 67 SNPs [4] (denoted as T1D67), all results are from that T1D GRS unless otherwise stated. As well as a linear set of terms it also includes a set of pair-wise interaction terms making the GRS non-linear as an overall function. The other two T1D GRSs consist of 10 (denoted T1D10) and 30 (denoted T1D30) [17] SNPs respectively; the Celiac GRS consists of 42 SNPs (denoted as CD42). All GRSs include pairwise-interaction terms making the overall GRS functions non-linear.

### 2.5 Statistical tests and metrics

We compared the masked-MLP approach to application of the standard GRSs on both uncorrupted and corrupted data. We used the test data and corrupted it in the same manner as when training the model; we randomly selected SNPs to corrupt with set probabilities, those that are selected are corrupted by alteration of the values uniformly to 0, 1, 2.

We corrupted the input data with masking probability of 0 (no corruption) to 0.25 (25% chance of any SNP being corrupted), in increments of 0.05. For example a masking probability during inference of 0.1 means that each SNP has a 10% change of being corrupted, with the corruption being altering the value to 0,1,2 with a uniform probability. Both the masked-MLP and standard GRS were run on all the corrupted data.

To explore the importance of the noise during training we trained the model with no input noise. We also trained models without the inclusion of proxy SNPs to assess the importance of those additional SNPs, i.e. the model trained with the original *p*_0_ SNPs. For those models we trained with the range of input noise levels from probabilities of 0 to 0.25 as for the models trained with proxies.

As metrics we considered the Spearman rank correlation coefficient between the estimates on the corrupted data (from the model or the GRS) and the GRS on the uncorrupted data. In addition, we showed the area under the receiver operating characteristic curve (AUC) between cases and population data. All confidence intervals were reported at the 95% level.

## 3 Results

In Figure 4 we show comparisons of the masked-MLP trained with a masking probability of 0.1 and the GRS for the equivalent level of masking probability during inference from 0 to 0.25. The masked-MLP was more robust to the introduction of noise than the GRS on the same input data. At a 2.5% masking percentage the GRS correlation and AUC fell to 0.889 (0.886-0.891) and 0.888 (0.870-0.905) respectively while the masked-MLP equivalent metrics were 0.970 (0.969-0.971) and 0.918 (0.903-0.931) respectively. The masked-MLP was robust to substantial levels of noise in the SNPs, when the masking probability is at 20% the correlation and AUC only fell to 0.909 (0.908-0.911) and 0.897 (0.880-0.912) respectively.

**Figure 4:**
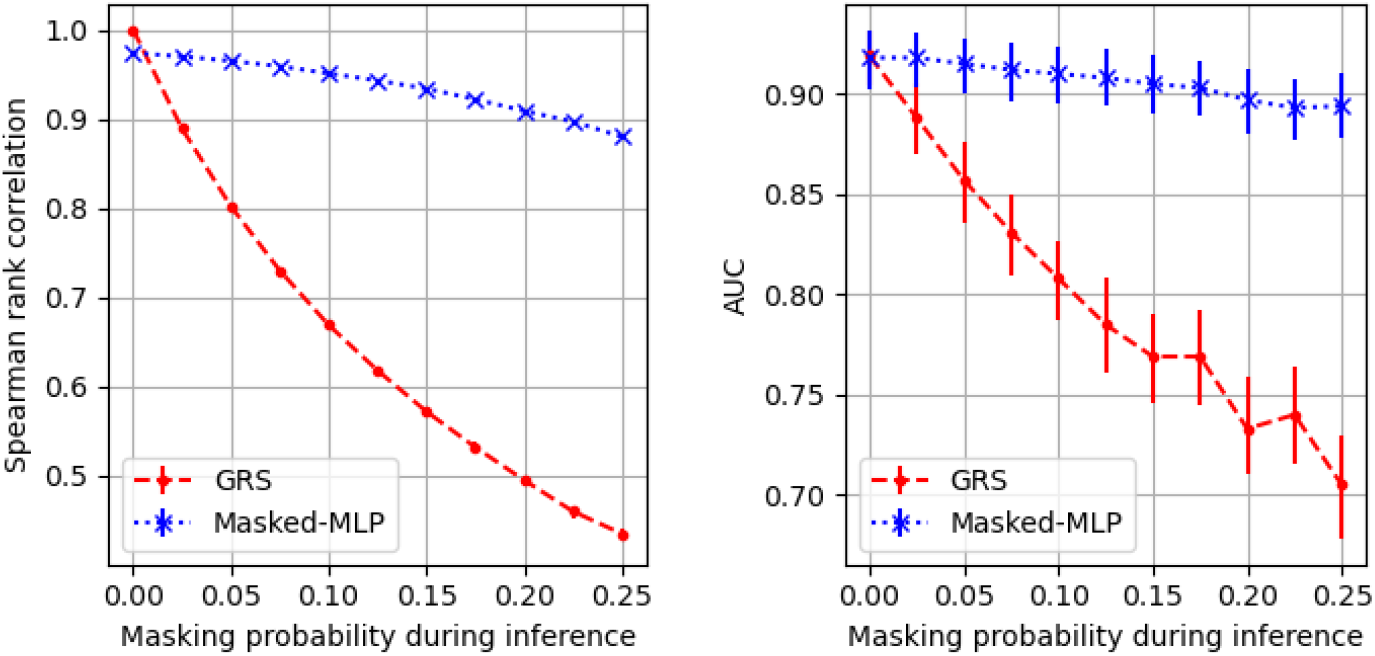
The Spearman rank correlation (left) and AUC (right) for the GRS and the masked-MLP model trained with a masking probability of 0.1. The masking probability during inference ranges from 0 to 0.25, each a different set of inputs.

The introduction of noise into the training process for the masked-MLP is important otherwise the model will not be robust to corruption when running inference (Figure 5). At an inference masking probability of 0.1 the model with no noise during training produces a correlation of 0.772 (0.769-0.775) with the original GRS and an AUC of 0.856 (0.839-0.875). Conversely, with a masking probability during training of 0.1 the correlation and AUC at an inference masking probability of 0.1 were at 0.951 (0.950-0.952) and 0.910 (0.895-0.924) respectively.

**Figure 5:**
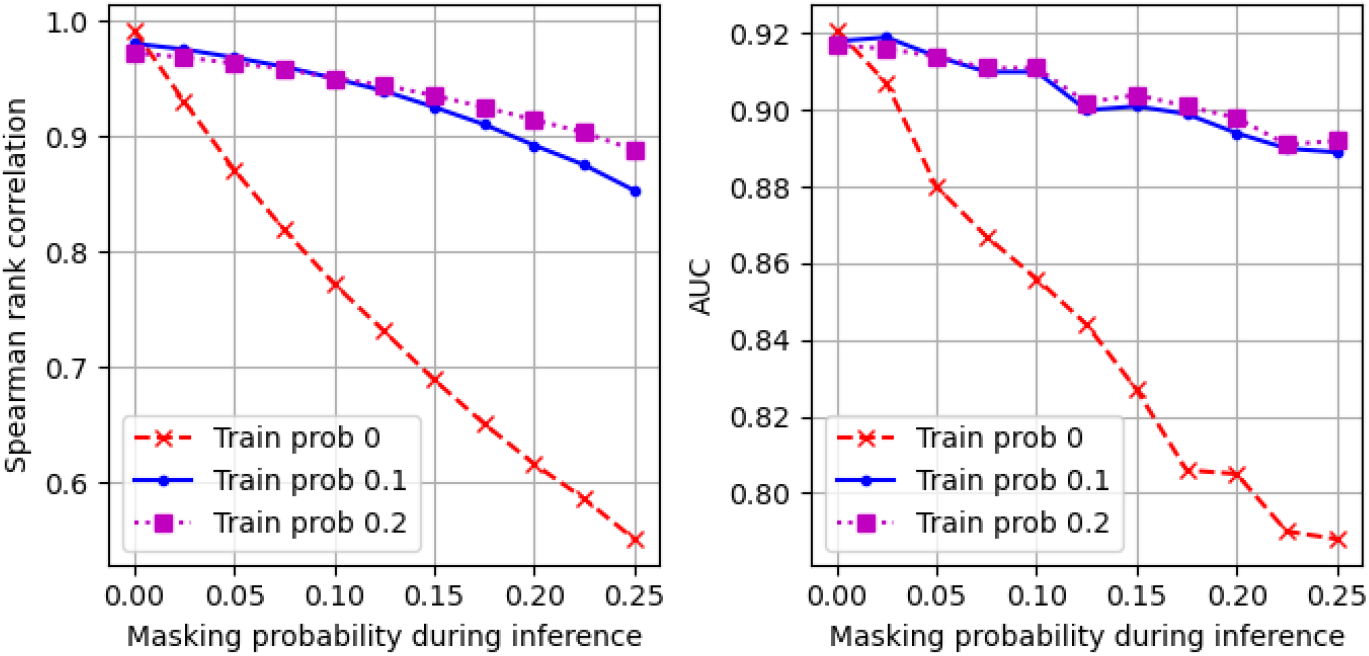
The importance of including noise into the training process. Results for the T1D67 GRS. Three sets of trained models are shown which were trained with masking probabilities of 0, 0.1 and 0.2 respectively (denoted as *Train prob*). Different levels of masking probability during inference are shown across the x-axis. Uncertainties are not displayed to make the plots clearer.

The effect on individual sample predictions of the inclusion of noise in the training data and inference data is shown in Figure 6. When noise is not added to the training data (left plot) the model produces predictions on the corrupted testing data that differ substantially from those on the uncorrupted testing data. Conversely, when the training data is corrupted (right plot) the model learns a representation that is more robust to noise, with more similar predictions on both corrupted and uncorrupted testing data.

**Figure 6:**
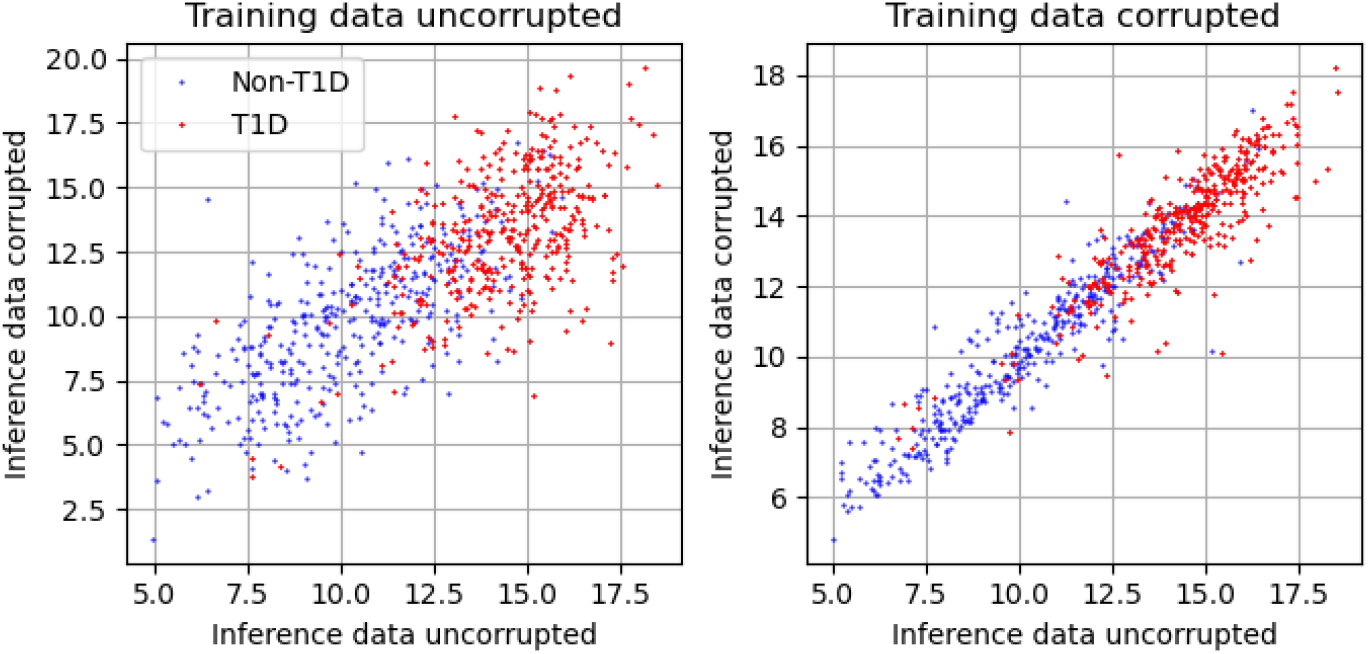
Sample-level model estimates made on both uncorrupted and corrupted testing data for models trained separately on uncorrupted and corrupted data. Participants with T1D (red dots) and non-T1D (blue dots) are shown with a random 387 non-T1D data-points selected. Left) T1D67 model predictions made on corrupted testing data versus uncorrupted testing data for a model trained on uncorrupted data. Right) T1D67 model predictions made on corrupted testing data versus uncorrupted testing data for a model trained on corrupted data at a probability of 0.1

Another key feature of the masked-MLP model is the inclusion of proxy SNPs (Figure 7). The model which is trained without any additional proxies shows a faster fall for both Spearman rank correlation (left plot) and AUC (right plot) than the model trained with proxies. At a level of inference noise (via masking probability) of 0.1 the model trained with no proxies had a Spearman rank correlation of 0.886 (0.884-0.888) compared to an equivalent for the model trained with proxies of 0.951 (0.950-0.952). For discrimination between cases and controls the AUC for the model with no proxies fell to 0.887 (0.872-0.902) at a masking probability during inference of 0.1 while the equivalent for the model with proxies had an AUC of 0.910 (0.895-0.924).

**Figure 7:**
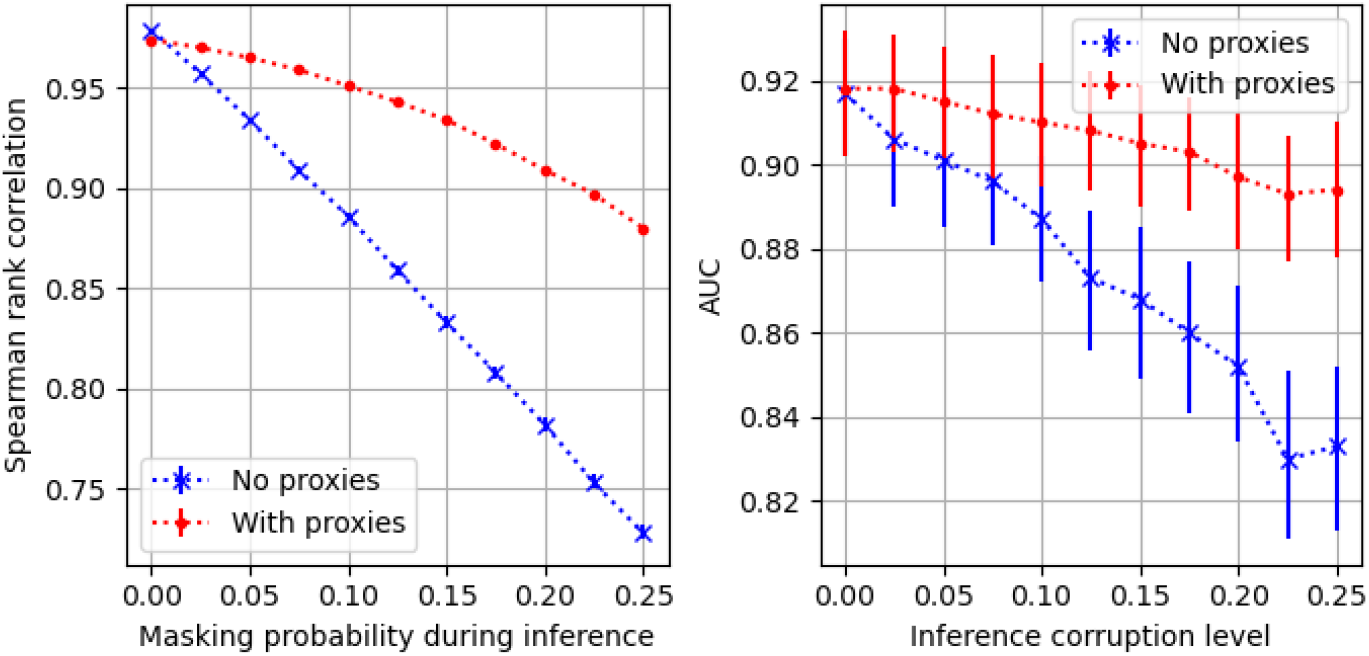
The effect of including proxies on the robustness of the masked-MLP. Results for a model with no additional proxy SNPs included (just the original 67 SNPs) are shown along with the standard masked-MLP with the addition of three proxy SNPs for each original SNP. Both models were trained with a masking probability of 0.1. Left) Spearman rank correlation. Right) AUC between cases and population samples.

In Figure 8 we show the masked-MLP and GRS results on corrupted data for the three other GRSs: T1D30, T1D10 and CD42. We show the Spearman rank correlation coefficient and AUC between cases and controls as for the other plots. The pattern remains the same for T1D30, T1D10 and CD42 as for the T1D67, with the masked-MLP being more robust than the GRS on the corrupted data.

**Figure 8:**
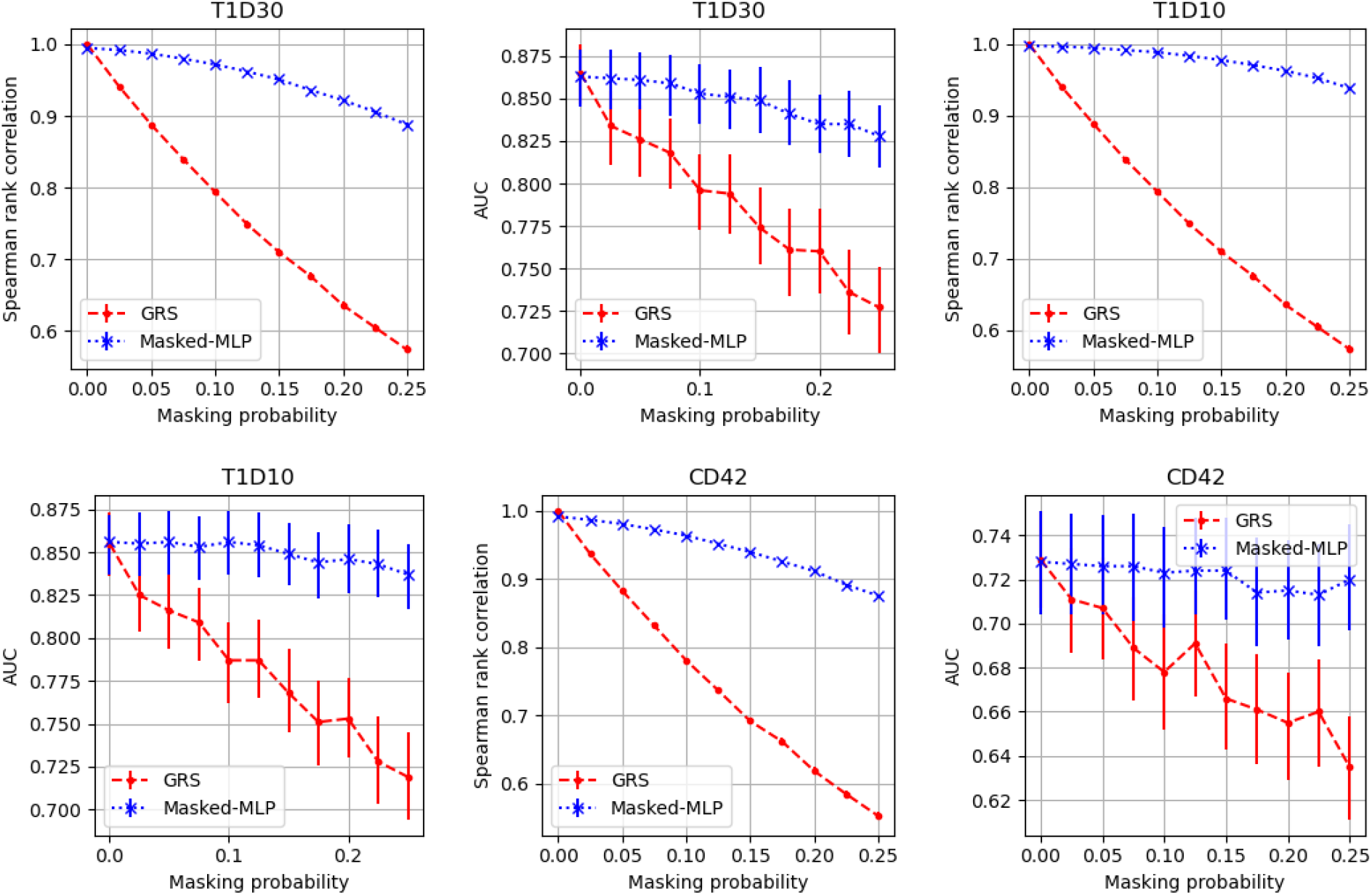
The Spearman rank correlation and AUC for the T1D30, T1D10 and CD42 GRSs and the associated masked-MLP models all trained with masking probability of 0.1. The masking probability during inference ranges from 0 to 0.25.

## 4 Discussion

GRSs can aid with predicting onset and progression of disorders or understanding traits in individuals. As GRSs are produced from multiple SNPs if any are missing, or not correctly estimated, the GRS value will be incorrect by some amount, dependent on the level of noise, the frequencies of the SNPs and their importance within the GRS. A wrongly calculated GRS could alter the risk profile of an individual, potentially resulting in missed opportunities to intervene for those with risk underestimated, or unnecessary additional interventions for those whose risk is overestimated. In addition, a poorly calculated GRS may lead to inaccurate conclusions being drawn, for example falsely inferring that the GRS is ineffective on a certain dataset. A more robust calculation of GRSs would reduce these effects.

Missing SNPs will usually reduce the quality of the GRS; either information is lost which reduces discrimination power, or if SNPs are replaced with proxies then, unless they are perfectly correlated, the SNP values will not be identical. The effect of the use of proxies is effectively the inclusion of additional noise into the GRS resulting in poorer quality; for a classification task this results in poorer discrimination performance. The impact of noise in the SNP arrays has a similar effect of reducing quality of the GRS. The overall impact is that the calculation of a standard GRS is sensitive to noise in the inputs or to the replacement of missing SNPs by proxies.

Our masked-MLP approach was more robust to noise than a standard GRS method. As the noise level of the testing data was increased the fall in both correlation to the original GRS and the AUC between cases and controls remained modest; which holds even for substantial levels of imposed noise.

Two key aspects of the masked-MLP that allow for a robust calculation of a GRS are the inclusion of additional proxy SNPs and the injection of noise during training. The additional proxy SNPs means the model has extra information to base its estimations on, allowing it to place less weight on one SNP and produce a more robust estimate. The addition of noise during training means the model cannot fully rely on the values of the individual SNPs and therefore is forced to learn a representation that assumes there will be corruption in the SNPs, leading to a more robust representation.

We demonstrated the importance of including noise during training by training models without the injection of noise. If no noise was added during training the model performed poorly once modest levels of corruption were added to the testing data, while those models with additional noise during training showed only a modest reduction in performance. The trade-off is that the addition of noise marginally reduces the quality of the GRS calculation if there is no noise in testing. However, the effects are small with no significant change for the AUC and only small reductions in correlation to the original GRS.

Similarly, without the addition of proxies the masked-MLP is not as robust. If only the original SNPs are included the model prediction quality (measured either as correlation with the original GRS or the AUC between cases and population samples) falls more substantially with increasing noise during inference than when proxies are included.

Our masked-MLP approach allows for a more robust GRS to be calculated. This has the potential to improve GRS calculation with benefits for both research and the use of GRSs in clinical settings.

## 5 Conclusion

GRSs are increasingly being developed and used for a variety of disorders. Standard methods of calculating a GRS can suffer from missing or noisy input data, resulting in potentially inaccurate and misleading risk estimates. Our approach uses a neural network architecture together with additional proxy SNPs and added noise in the training procedure to produce a more robust method of calculating GRSs.

## Data Availability

UK Biobank data is available to researchers through application to: //www.ukbiobank.ac.uk/.

